# Cerebrospinal fluid A beta 1-40 peptides increase in Alzheimer’s disease and are highly correlated with phospho-tau in control individuals

**DOI:** 10.1101/2020.06.02.20119578

**Authors:** Sylvain Lehmann, Julien Dumurgier, Xavier Ayrignac, Cecilia Marelli, Daniel Alcolea, Juan Fortea Ormaechea, Eric Thouvenot, Constance Delaby, Christophe Hirtz, Jérôme Vialaret, Nelly Ginestet, Elodie Bouaziz-Amar, Jean-Louis Laplanche, Pierre Labauge, Claire Paquet, Alberto Lleo, Audrey Gabelle, for the Alzheimer’s Disease Neuroimaging Initiative (ADNI)

**Author notes:** **Corresponding author:** Sylvain Lehmann. Data used in preparation of this article were obtained from the Alzheimer’s Disease Neuroimaging Initiative (ADNI) database (adni.loni.usc.edu). As such, the investigators within the ADNI contributed to the design and implementation of ADNI and/or provided data but did not participate in analysis or writing of this report. A complete listing of ADNI investigators can be found at: http://adni.loni.usc.edu/wpcontent/uploads/how_to_apply/ADNI_Acknowledgement_List.pdf.

## Abstract

**Background:** Amyloid pathology, which is one of the characteristics of Alzheimer’s disease (AD), results from altered metabolism of the beta-amyloid peptide (Aβ) in terms of synthesis, clearance or aggregation. A decrease in cerebrospinal fluid (CSF) level Aβ 1-42 is evident in AD, and the CSF ratio Aβ 40 /Aβ 40 has recently been identified as one of the most reliable diagnostic biomarkers of amyloid pathology. Variations in inter-individual levels of Aβ 1-40 in the CSF have been observed in the past, but their origins remain unclear. In addition, the variation of Aβ 40 in the context of AD studied in several studies has yielded conflicting results.

**Methods:** Here, we analyzed the levels of Aβ 1-40 using multicenter data obtained on 2466 samples from six different cohorts in which CSF was collected under standardized protocols, centrifugation and storage conditions. Tau and p-tau(181) concentrations were measured using commercially available in vitro diagnostic immunoassays. Concentrations of CSF Aβ 1-42 and Aβ 1-40 were measured by ELISA, xMAP technology, chemiluminescence immunoassay (CLIA) and mass spectrometry. Statistical analyses were calculated for parametric and non-parametric comparisons, linear regression, correlation and odds ratios. The statistical tests were adjusted for the effects of covariates (age, in particular).

**Results:** Regardless of the analysis method used and the cohorts, a slight but significant age-independent increase in the levels of Aβ 40 in CSF was observed in AD. We also found a strong positive correlation between the levels of Aβ 40 and p-tau(181) in CSF, particularly in control patients.

**Conclusions:** These results indicate that an increase in the baseline level of amyloid peptides, which are associated with an increase in p-tau(181), may be a biological characteristic of AD. This confirms the potential therapeutic value of lowering the baseline levels of Aβ 40 which, being elevated, can be considered a risk factor for the disease.

## Background

Alzheimer’s disease (AD) neuropathological brain lesions consist of aggregates of hyperphosphorylated tau proteins, which have also been called neurofibrillary tangles (NFTs), and extracellular deposits of amyloid precursor protein (APP) derived amyloid-beta peptides (Aβ), which are known as amyloid plaques. Much research has been focusing recently on the molecular mechanisms underlying these pathological events as it has become essential to develop preventive and therapeutic strategies for AD. For a long time, the main explanation for the pathogenesis of AD was that amyloidogenesis was the *primum movens* of the affection, which led to the concept of the amyloid cascade [1]. According to this picture of the disease, the alteration of APP metabolism (increasing amyloid production, decreasing clearance rates..), the aggregation of Aβ peptides and the formation of amyloid plaques, might result in microglial and astrocyte activation, local inflammatory responses, oxidative stress and eventually in the hyperphosphorylation of tau proteins and secondarily, in the formation of NFTs [2].

The idea that amyloid peptides contribute importantly to the etiology of AD is supported by cases of AD who carry presenilins (1 or 2) or APP mutations [3]. These gene mutations trigger the overproduction of Aβ peptides or the preferential production of Aβ42, which is the most amyloidogenic of all the peptides. An APP gene dose effect triggering AD development, as occurs in Down syndrome [4] and in gene duplication processes [5], is a further/an additional potential factor contributing to amyloid pathogenesis. Other genetic factors have been described, such as apolipoprotein E4 allele, in particular [6].

Studies on cerebrospinal fluid (CSF) biomarkers in AD have greatly improved our understanding of the pathophysiology of this disease. The production of amyloid peptides following the neuronal processing of APP has been involved in the response to physiological challenge with neurotrophic, anti-microbial, tumor suppression or synaptic function regulation activities [7].

Regarding tau proteins which are associated to microtubules, their physiological secretion by neuronal cells is a recent discovery which physiological relevance and benefit is still matter of debate [8]. A decrease in CSF Aβ 42 is especially indicative of an amyloidogenic process, while an increase in tau proteins (total tau and its phosphorylated form p-tau(181)) is known to be associated with axonal loss and tau pathology in AD [9, 10]. Tests on these two biomarkers are being included nowadays in the international clinical research guidelines [11, 12], and many centers [11–15], and ourselves [13–15] have integrated them into daily clinical practice. Importantly, these biochemical CSF measurements are concordant with the results of the PET imaging approaches which were initially developed to determine the brain amyloid load [16], and now also serve to measure tau accumulation [17]. These data are in line with hypotheses put forward by Jack *et al*, [18] about the chronology of the evolution of biomarkers during the pathophysiological process, and the relevance of amyloid markers in particular at a very early stage, probably as early as 10 to 15 years before the onset of clinical symptoms.

Under non-pathological conditions, Aβ40 is highly correlated with Aβ42 [19]. The computation of the ratio Aβ42/40 is now being used in routine clinical practice on AD patients in some centers [20–22]. This is a useful approach for reducing pre-analytical Aβ42 biases [23–25] and improving the diagnostic performances of CSF biomarkers [26], especially in discordant cases [27]. This ratio can also be used to account for interindividual amyloid variations in the baseline CSF level [28]. Low CSF Aβ40 levels might also be indicative of frontotemporal dementia (FTD) [29, 30], cerebral amyloid angiopathy (CAA) [31], HIV [32], multiple sclerosis [33] or normal pressure hydrocephalus [34].

In their meta-analysis of AD biomarkers, Olsson et al. [35] observed the existence of a negligible difference in CSF Aβ40 between AD and control patients. Most of the 32 studies considered had a limited number of patients included in each group (the median number of subjects per group was less than 30, and the maximum number of subjects was 137 and 328 in AD and non-AD groups, respectively). The focus of these studies were also quite different, looking at the diagnostic interest of Aβ42/40 in AD, of of Aβ peptides in other neurodegenerative diseases, or being more interested in pathophysiological mechanisms. In the present study, we revisited the issue of the Aβ40 levels using large series of multicentre data. The results show the occurrence of a significant age-independent increase in CSF Aβ40 in AD. Another noteworthy finding was the existence of a strong positive correlation between CSF Aβ40 and the p-tau(181) concentration, even in patients without Alzheimer’s disease (NAD). These findings suggest that the baseline amyloid peptide level may constitute a risk factor contributing to sporadic AD, which is associated with p-tau(181) production.

## Methods

### Study design and subjects

Patients with cognitive impairments were recruited and followed at the Montpellier and Paris Memory Resources Center (CMRR). The Montpellier participants were subdivided into two cohorts which were recruited during different periods: Montpellier 1 (Mtp-1) (recruited from 07/2015 to 05/2017) and Montpellier 2 (Mtp-2) (recruited from 09/2009 to 06/2015). These two periods corresponded to the use of different ELISA kits (SupTable 1). The cohort Mtp-1 consisted of 400 patients (126 AD, 274 NAD), the cohort Mtp-2 consisted of 504 patients (220 AD, 284 NAD). The Paris cohort consisted of 624 patients (299 AD, 325 NAD) from the Centre de Neurologie Cognitive, Groupe Hospitalier Lariboisière Fernand-Widal (recruited from 03/2012 to 05/2017). The Barcelona SPIN (Sant Pau Initiative on Neurodegeneration) cohort (79 AD, 148 NAD) consisted of patients who had undergone lumbar puncture for CSF AD biomarkers at the Sant Pau Memory Unit [36, 37] (recruited from 05/2009 to 12/2017). All the patients underwent a thorough clinical examination including biological lab tests, neuropsychological assessments and brain imaging. The same diagnostic procedure [27] and AD diagnostic criteria [38] were used at all the clinical centres which participated. The NAD diagnosis included FTD based on relevant criteria [39], dementia with Lewy bodies based on the McKeith criteria [40]), corticobasal degeneration (based on the criteria defined by Boeve et al [41]), progressive supranuclear palsy, amyotrophic lateral sclerosis, chronic hydrocephalus, vascular dementia and psychiatric disorders (based on the usual consensus diagnostic criteria). All the patients at each clinical centre gave their written informed consent to participating in clinical research on CSF biomarkers, which was approved by the respective Ethics Committees. The committee responsible in Montpellier was the regional Ethics Committee of the Montpellier University Hospital and Montpellier CSF-Neurobank #DC-2008-417 at the certified NFS 96-900 CHU resource center BB-0033-00031, www.biobanques.eu. Authorization to handle personal data was granted by the French Data Protection Authority (CNIL) under the number 1709743 v0. Two sets of data originating from the analysis of CSF samples at the Alzheimer’s Disease Neuroimaging Initiative (ADNI) database (www.loni.ucla.edu/ADNI) were used after the agreement of the scientific committee. ADNI UPEN-RESULTS, UPEN-ELYCYS (n = 311) and MS UPENNMSMSABETA (n = 400) data were also used. In the ADNI cohorts, which included many patients with mild cognitive impairments (MCI), we had to rely on the biological PLM (Paris-Lille-Montpellier) scale [42] to define populations with a low (ADNI(-)) and high (ADNI(+)) prevalence of AD. This scale combines the concentration of the three CSF biomarkers [Aβ42, tau, ptau(181)] into a probability scale for AD. The score ranges from 0 to 3 based on the number of abnormal CSF biomarkers. ADNI(-) population corresponded to PLM scale of 0 or 1 with less than 25% of AD, while ADNI(+) corresponded to PLM scale of 2 and 3 with more than 75% of AD. Importantly the PLM score used was not based on the Aβ40 values so as to prevent circular reasoning. This way of stratifying patients in the ADNI cohort represents anyway a limitation of our study.

### CSF samples and assays

CSF was collected using standard conditions of collection, centrifugation and storage [43, 44]. CSF tau and p-tau(181) concentrations were measured using the standardized commercially available INNOTEST_R_ sandwich ELISA, Luminex® xMAP technology (x = analyte, MAP = Multi-Analyte Profiling) assays in line with the manufacturer’s instructions (Fujirebio-Europe). The consistency of the p-tau(181) detection using the ELISA assays is ensured by its comparison with the mass spectrometry detection performed in this fluid [45]. In the Mtp-1 cohort, CSF Aβ1-42 and Aβ1-40 (denoted here by Aβ42 and Aβ40) were measured with Euroimmun kits (EQ-6511-9601 (Aβ1-40); EQ-6521-9601 (Aβ1-42)). In the Mtp-2 and Paris cohorts, CSF Aβ42 and Aβ40 were measured using INNOTEST_R_ sandwich ELISA from IBL and Fujirebio, respectively, as recommended by the manufacturer. Roche Elecsys automated chemiluminescence immunoassay (CLIA) and mass spectrometry were used on the ADNI cohorts to measure CSF Aβ1-42 and Aβ1-40 as previously described [46, 47]. Detection limits of these kits are compatible with CSF clinical ranges. Average concentration of analytes may differs between kits in relation with standard value assignments by the vendors in the absence of certified reference materials.

The pre-analytical procedure was standardized [44] but differed, depending on the type of collection tubes used [36, 48]). This explains the differences observed between cohorts in the mean aβ40 and Aβ42 values measured with the same detection kit (SupTable 1). The quality of the results was ensured by using validated standard operating procedures and internal quality controls (QCs). The QC coefficient of variation obtained on the CSF analytes in each batch and between batches ranged consistently below 15%. In addition, external QC procedures were used to confirm the quality/accuracy of the results [43]. In the case of the ADNI cohorts, CSF samples were deep frozen after the lumbar puncture without performing any centrifugation or aliquoting, and shipped to the UPENN ADNI Biomarker Laboratory in Philadelphia on dry ice, where they were thawed, aliquoted, and re-frozen.

### Statistical analysis

Statistical analyses were computed with the MedCalc software program (18.11.3). Data tested for normality are expressed as means ± SDs, and differences between groups were taken to be significant in the Student’s t-tests at P < 0.05. Linear regression was computed between continuous biomarkers, and the corresponding Pearson correlation coefficients and statistical significance have been specified in the tables. When indicated, statistical tests were adjusted to account for the effects of covariates (age, in particular). Odds ratios corresponded to the presence of AD in the various percentile groups, based on the distribution of Aβ40. The 95% confidence odds ratio intervals were computed along with the z-statistics and the associated P-values.

## Results

### CSF Aβ42 and Aβ40 in AD and NAD populations

CSF data on 2466 samples originating from six different cohorts were included in the present study. The AD and NAD populations were defined based on clinical criteria in the Montpellier 1 (Mtp-1), Montpellier 2 (Mtp-2), Paris and Barcelona cohorts, and on the PLM score for the ADNI cohorts. Differences were observed in terms of age and CSF biomarker profiles in the overall population, as well as in each clinical cohort (Table 1). As was to be expected, AD patients obtained lower MMSE scores and showed higher CSF tau and p-tau(181) levels than NAD patients/participants. A significant decrease in the CSF Aβ42 concentrations was observed in the AD population (Figure 1A), regardless of the cohort tested. Noteworthy differences were also observed in the Aβ40 levels between all the cohorts (Figure 1B): the values recorded in the AD population were significantly higher than in the NAD group regardless of analytical method, the sex or the age as covariate.

**Figure 1:**
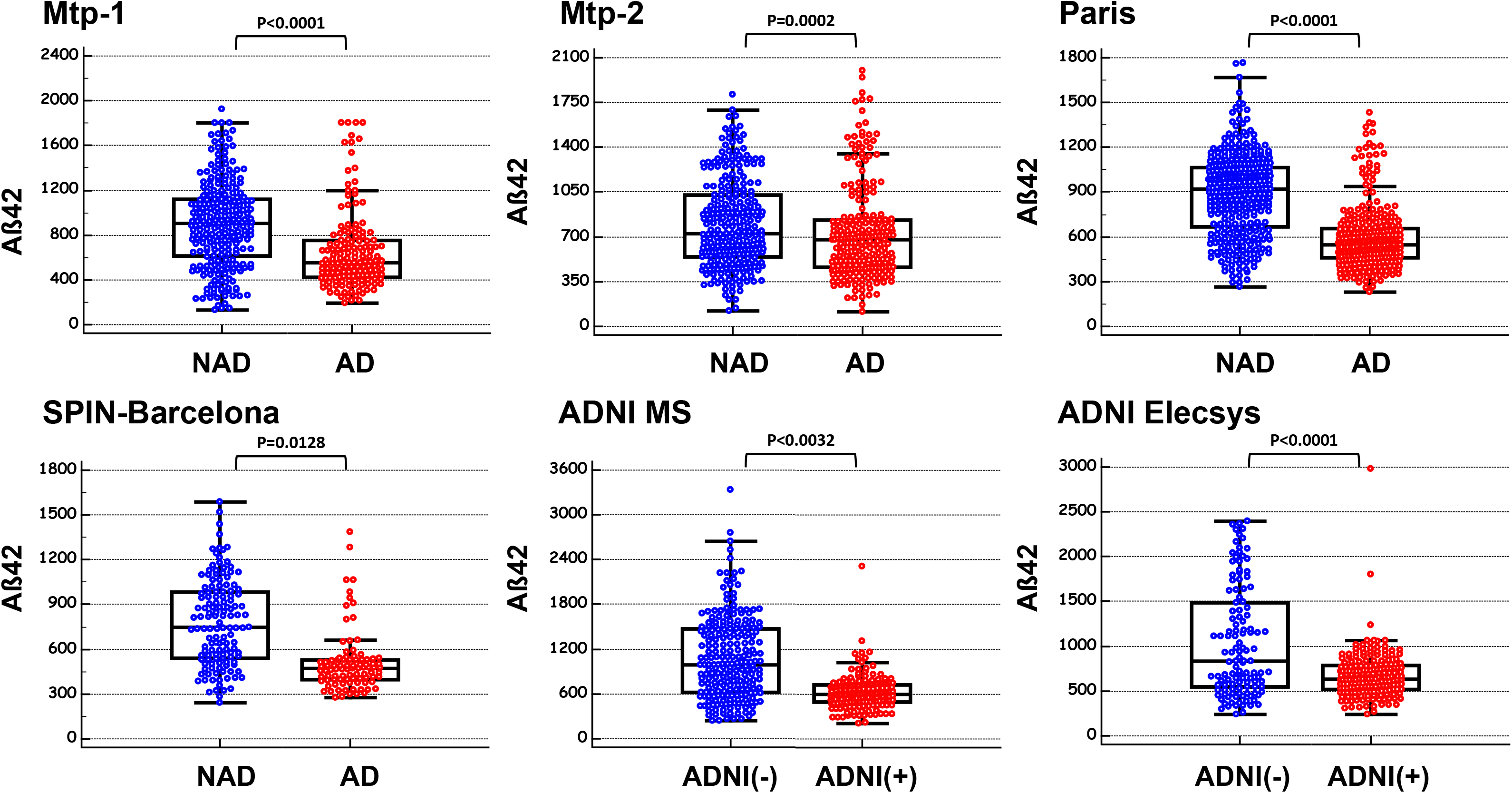
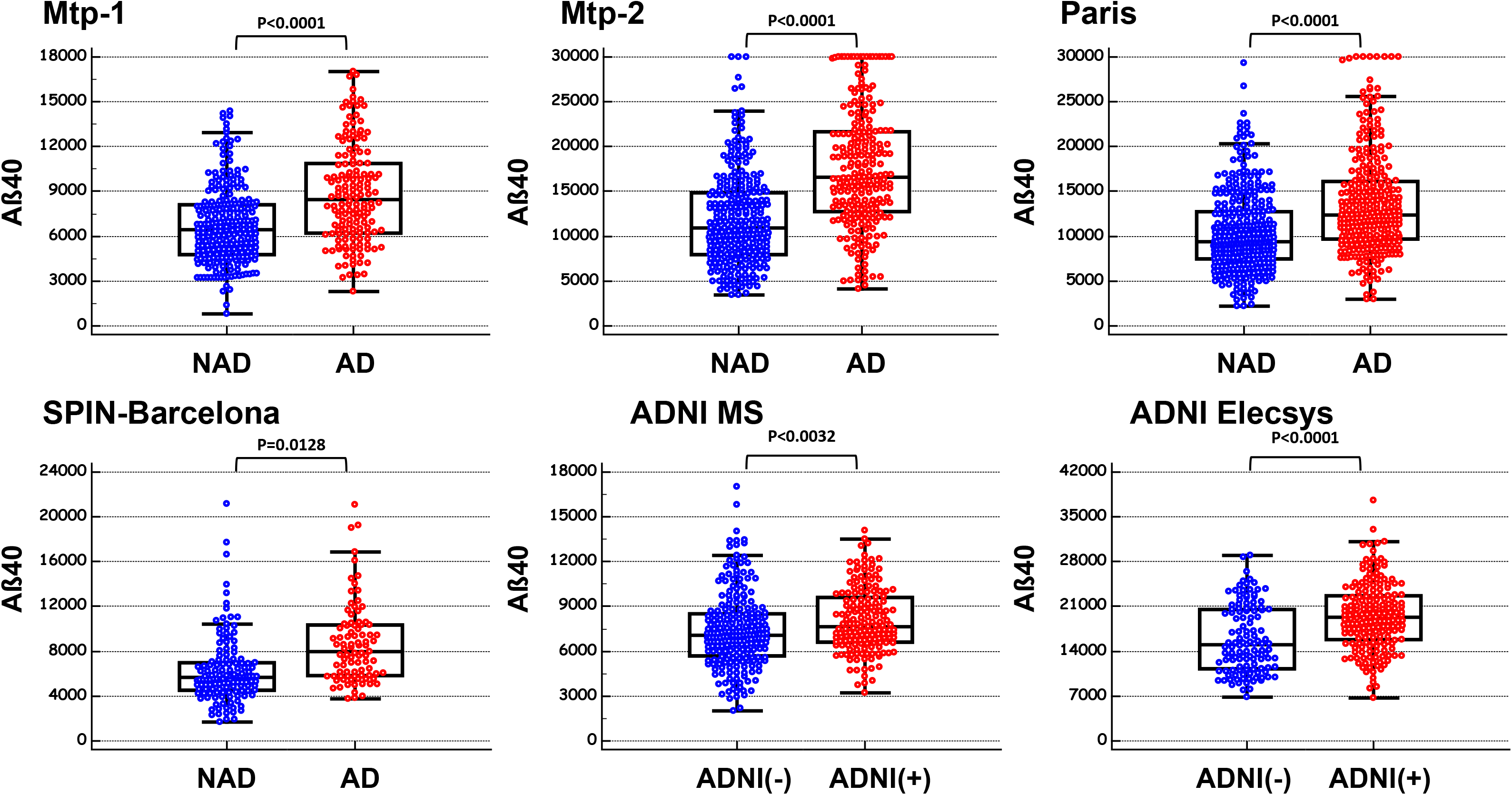
CSF Aβ42 and Aβ40 in Non-AD and AD populations. CSF concentration of Aβ42 (panel A) and Aβ40 (panel B) in six independent cohorts (Montpellier 1 (Mtp-1), Montpellier 2 (Mtp-2), Paris, SPIN-Barcelona, ADNI-MA, ADNIElecsys) confirmed the significant difference between NAD and AD patients for both analytes (ttest). Note that Aβ has been assess using five different detection methods (supTable 1).

**Table 1.** Demographical and cerebrospinal fluid (CSF) biomarkers characteristics of the six cohorts, Montpellier 1 (Mtp-1), Montpellier 2 (Mtp-2), Paris, SPIN-Barcelona, ADNI-MS and ADNIElecsys. Results are expressed as the mean +/− standard deviation (SD). Abbreviation: MMSE = mini mental state examination; AD = Alzheimer’s disease; NAD = Non Alzheimer’s disease; ADNI(-) = cognitive patients with non Alzheimer’s disease PLM profile; ADNI(+) = cognitive patients with Alzheimer’s disease PLM profile; P significance level of the Student’s t-test and * Chi-squared test for the comparison of two proportions. Values of Aβ40, Aβ42, tau and p-tau(181) are in pg/mL.

| Cohort / analyte | NAD |  | AD |  |  | Cohort / analyte | NAD |  | AD |  |  |
| --- | --- | --- | --- | --- | --- | --- | --- | --- | --- | --- | --- |
| Mtp-1 | Mean | SD | Mean | SD | P | SPIN-Barcelona | Mean | SD | Mean | SD | P |
| Aβ40 | 7035 | 2800 | 8579 | 3274 | <0.0001 | Aβ40 | 6799 | 3633 | 7985 | 2891 | 0.0128 |
| Aβ42 | 909 | 399 | 561 | 237 | <0.0001 | Aβ42 | 793 | 288 | 440 | 81 | <0.0001 |
| TAU | 343 | 279 | 739 | 267 | <0.0001 | TAU | 284 | 223 | 727 | 392 | <0.0001 |
| PTAU | 42 | 16 | 96 | 36 | <0.0001 | PTAU | 47 | 22 | 103 | 47 | <0.0001 |
| Age | 67.0 | 11.4 | 70.7 | 9.2 | 0.0015 | Age | 66.1 | 10.1 | 70.8 | 8.3 | 0.0004 |
| MMSE | 22.1 | 6.2 | 19.8 | 7.5 | 0.1392 | MMSE | 26.6 | 6.2 | 23.5 | 4.6 | 0.0097 |
| Sex (%M) | 53.6% | / | 38.1% | / | 0.0040* | Sex (%M) | 57.4% | / | 38.0% | / | 0.0055* |
| ApoE (% E4) | NA |  |  |  |  | ApoE (% E4) | 63.3% | / | 49.4% | / | 0.1910* |
| Cohort / analyte | NAD |  | AD |  |  | Cohort / analyte | ADNI(-) |  | ADNI(+) |  |  |
| Mtp-2 | Mean | SD | Mean | SD | P | ADNI-MS | Mean | SD | Mean | SD | P |
| Aβ40 | 12720 | 6046 | 16802 | 6473 | <0.0001 | Aβ40 | 7387 | 2517 | 8101 | 2167 | 0.0032 |
| Aβ42 | 821 | 369 | 700 | 334 | 0.0002 | Aβ42 | 1076 | 547 | 620 | 231 | <0.0001 |
| TAU | 321 | 248 | 655 | 302 | <0.0001 | TAU | 66 | 24 | 140 | 58 | <0.0001 |
| PTAU | 42 | 18 | 89 | 37 | <0.0001 | PTAU | 23 | 8 | 50 | 18 | <0.0001 |
| Age | 66.0 | 12.7 | 70.0 | 9.3 | 0.0001 | Age | 75.4 | 6.7 | 74.0 | 7.5 | 0.0430 |
| MMSE | 21.9 | 7.3 | 19.6 | 5.4 | 0.0031 | MMSE | 27.3 | 2.4 | 25.9 | 2.6 | <0.0001 |
| Sex (%M) | 51.4% | / | 50.9% | / | 0.9114* | Sex (%M) | 61.0% | / | 58.6% | / | 0.6349* |
| ApoE (% E4) | ND |  |  |  |  | ApoE (% E4) | 53.8% | / | 89.3% | / | <0.001* |
| Cohort / analyte | NAD |  | AD |  |  | Cohort / analyte | ADNI(-) |  | ADNI(+) |  |  |
| Paris | Mean | SD | Mean | SD | P | ADNI-Elecsys | Mean | SD | Mean | SD | P |
| Aβ40 | 10767 | 4468 | 13149 | 5850 | <0.0001 | Aβ40 | 15865 | 5265 | 19417 | 5224 | <0.0001 |
| Aβ42 | 925 | 277 | 574 | 200 | <0.0001 | Aβ42 | 1039 | 610 | 673 | 269 | <0.0001 |
| TAU | 236 | 134 | 609 | 280 | <0.0001 | TAU | 241 | 68 | 433 | 148 | <0.0001 |
| PTAU | 42 | 16 | 90 | 37 | <0.0001 | PTAU | 21 | 6 | 45 | 18 | <0.0001 |
| Age | 65.2 | 9.4 | 70.7 | 8.1 | <0.0001 | Age | 72.9 | 7.3 | 72.8 | 6.3 | 0.9155 |
| MMSE | 23.0 | 5.2 | 21.8 | 5.8 | 0.0069 | MMSE | 27.1 | 2.4 | 25.4 | 3.6 | <0.0001 |
| Sex (%M) | 51.9% | / | 38.5% | / | 0.0008* | Sex (%M) | 59.8% | / | 49.7% | / | 0.0816* |
| ApoE (% E4) | NA |  |  |  |  | ApoE (% E4) | 49.7% | / | 38.6% | / | 0.1538* |

In the cohorts that combined AD, MCI, FTD, Control (subjective cognitive impairment) and other neurological diseases (OND) patients, the difference between AD and the other clinical groups was eventually confirmed (Figure 2A). The influence of *APOE* status, which was available in the case of 983 samples (524 NAD with 36.6% E4+; 459 AD with 58.4% E4+), was also assessed with respect to the Aβ levels (SupFigure 1A-F). As previously reported [49, 50], the presence of ApoE4 was significantly associated with lower Aβ42 levels, as well as lower Aβ40 levels. The difference in Aβ40 values between NAD and AD patients was also observed in both ApoE4 positive and negative populations (SupFigure 1G-I).

**Figure 2:**
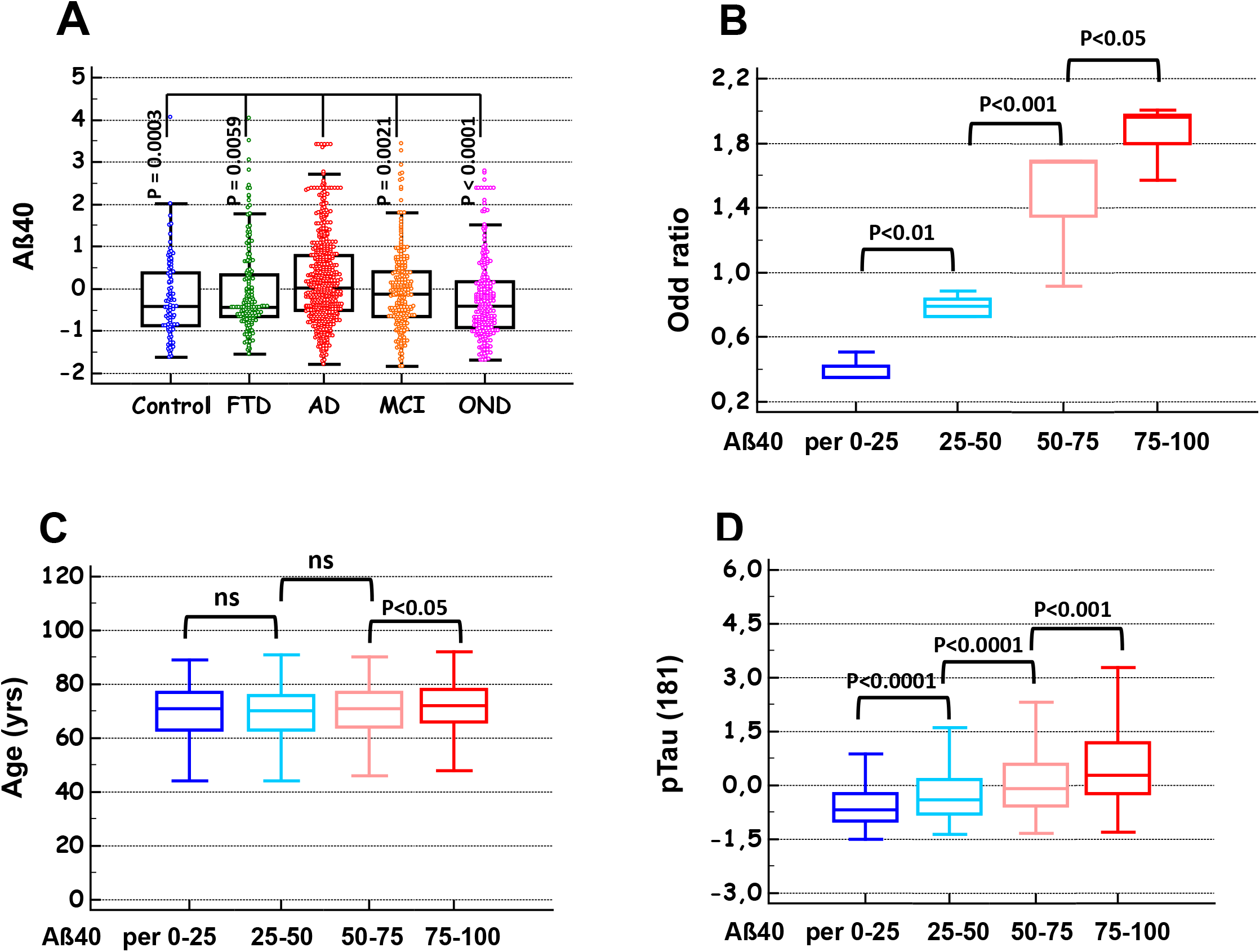
Aβ40 in different diagnosis; Representation in percentile; AD odd ratio, age and p-tau(181) distribution. The SPIN-Barcelona, Mtp-2 and Paris cohorts displayed a large range of pathological samples from patients with AD, Mild Cognitive Impairment (MCI), FTD, Control (subjective cognitive impairment) and other neurological diseases (OND). Mean-centred Aβ40 values in these cohorts were combined and compared in the different clinical groups confirming the significant increase of the peptides in AD (Panel A). The four cohorts (Montpellier 1 (Mtp-1), Montpellier 2 (Mtp-2), Paris, SPIN-Barcelona) have been sorted in four classes based on their Aβ40 percentile values as follows; p25: < 25^th^ percentile, p25-50: 25^th^-50^th^ percentile; p50-75: 50^th^-75^th^ percentile; p75: > 75^th^. The odd ratio for AD (panel B), the age of the patients (panel C) and the concentration of CSF p-tau(181) (panel D) were then plotted in each percentile class. Significant differences between classes are indicated.

To investigate more closely the relationship between Aβ40 and AD diagnosis, the total population was sorted into four percentile classes based on the value of this biomarker in the CSF (< 25th, 25th-50th, 50th-75th and > 75^th^ percentiles). The percentage of AD patients in each cohort clearly increased along with the Aβ40 percentiles (SupTable2). To account for the differences in AD prevalence between the cohorts, the odds ratios for AD were plotted in the case of increasing Aβ40 percentile classes, and a significant increase ranging from 0.4 to 1.8 was observed (Figure 2B). To establish whether the difference in age observed between NAD and AD patients (SupTable 1) might be a significant determinant here, the age distribution between percentile classes was also plotted (Figure 2B). A significant difference in age distribution was observed only between the 50^th^/75th and the > 75^th^ Aβ40 percentile classes (Figure 2C). Age cannot therefore account for the association between Aβ40 levels and AD prevalence. Nevertheless further statistical test were adjusted using age as covariate.

### Correlations between Aβ40 and the other CSF biomarkers

The correlation between Aβ40 and the other CSF biomarkers was computed in global, NAD and AD populations, for each cohort, and in the overall population (Table 2). As was to be expected [19], a correlation was found to exist between Aβ40 and Aβ42, especially in the NAD group. Aβ40 was also correlated with the tau levels, and it was striking that the highest correlation coefficients were obtained with p-tau (181) rather than with t-tau, especially in the Mtp-1 cohort (a significant difference was observed between the correlation coefficients at P = 0.02). The correlation was clearly visible when the mean-centred p-tau(181) values were plotted in the various Aβ40 percentile classes (Figure 2D), showing significant differences between classes. This correlation had to be put in perspective with the fact that both analytes increased in AD with the patients’age (SupTable 1), which justifies the adjustments made for age in our statistical analysis. We illustrated graphically the correlations in the NAD and the AD populations which had much higher and more widely distributed p-tau(181) values (Figure 3AB). Interestingly, the correlation coefficients were maintained and became even higher in the NAD population. To document the relationship between Aβ40 and p-tau(181) outside the context of AD, these correlation coefficients were tested in a series of clinically defined patients with multiple sclerosis [51] and FTD [52] (Figure 3CD). The corresponding correlation coefficients were both significantly higher in these groups than in the AD population (P< 0.001).

**Figure 3:**
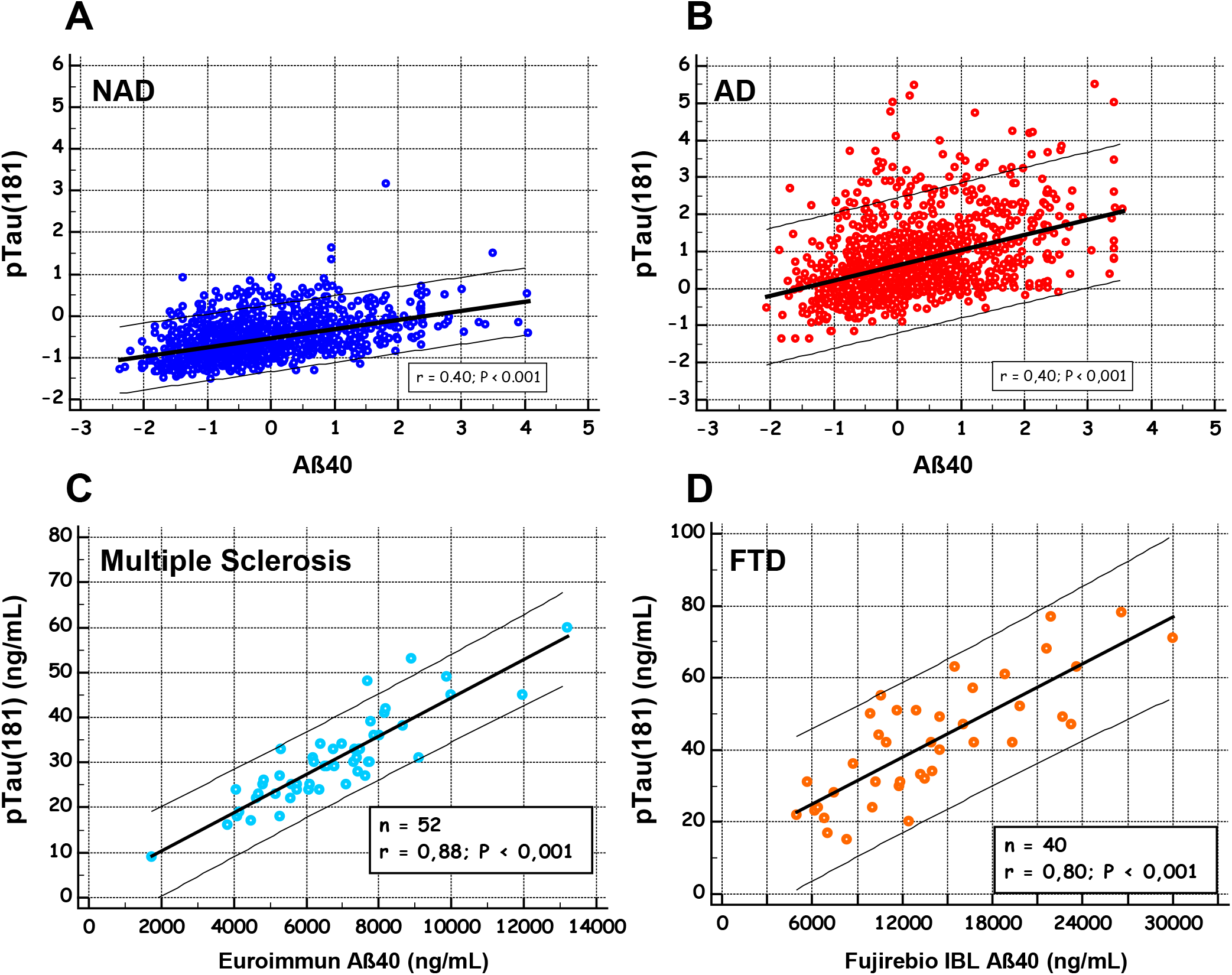
Correlation between Aβ40 and p-tau(181) in different clinical populations. To illustrate the correlation between Aβ40 and p-tau(181) (table 2), the mean-centered concentrations of the two analytes in the total study population were plotted in NAD (panel A) and AD populations (panel B). Aβ40 and p-tau(181) concentration were also plotted in a selection of multiple sclerosis (panel C) and FTD patients (panel D).

**Table 2.** Age adjusted Pearson correlation between Aβ40 and Aβ42, tau or p-tau(181) values in the six cohorts (Montpellier 1 (Mtp-1), Montpellier 2 (Mtp-2), Paris, SPIN-Barcelona, ADNI-MS, ADNI-Elecsys), in the overall population using mean-centred values to account for level differences between analytical methods. Computation has been done in the All population and in the NAD, AD, ADNI(-), ADNI(+) groups. Correlation coefficient statistical value P< 0.001 for all but * P< 0.01.

| Aβ40 | All | | | NAD or ADNI(-) | | | AD or ADNI(+) | | |
| --- | --- | --- | --- | --- | --- | --- | --- | --- | --- |
| correlation | pTau | Tau | Aβ42 | pTau | Tau | Aβ42 | pTau | Tau | Aβ42 |
| **Mtp-1** | 0.501 | 0.377 | 0.520 | 0.557 | 0.235* | 0.764 | 0.504 | 0.443 | 0.613 |
| **Mtp-2** | 0.514 | 0.443 | 0.407 | 0.430 | 0.244 | 0.533 | 0.304 | 0.213* | 0.469 |
| **Paris** | 0.419 | 0.36 | 0.188 | 0.421 | 0.145* | 0.571 | 0.288 | 0.245 | 0.247 |
| **SPIN-Barcelona** | 0.468 | 0.396 | 0.161 | 0.483 | 0.348 | 0.419 | 0.355 | 0.256* | 0.308* |
| **ADNI_MS** | 0.254 | 0.413 | 0.505 | 0.257 | 0.496 | 0.705 | 0.223* | 0.470 | 0.574 |
| **ADNI_Elecsys** | 0.536 | 0.567 | 0.449 | 0.487 | 0.624 | 0.782 | 0.502 | 0.496 | 0.580 |
| **Overall** | 0.445 | 0.418 | 0.368 | 0.455 | 0.318 | 0.615 | 0.339 | 0.318 | 0.426 |

## Discussion

The decrease in Aβ42 observed in the CSF of AD patients has attracted considerable attention in clinical and research communities. This decrease is attributable to the accumulation of Aβ42 in the brain parenchyma, along with a decrease in the rates of CSF clearance and an increase in the production of oligomeric/multimeric forms. Since determining the CSF Aβ42/Aβ40 ratio provides a useful means of improving AD diagnosis, many groups are now also measuring Aβ40 in their patients. A meta-analysis was however not conclusive regarding its differential levels in AD [35]. Looking back in detail at various studies, Aβ40 was either lower or showed no significant changes [21, 29, 30, 53], or apparently increased in AD in comparison with other forms of dementia [54, 55]. In a recent report, the increase in Aβ40 was clearly identified as one of the reasons for the good performances of the Aβ42/Aβ40 ratio as an index [20], while another study on PET amyloid findings also established that CSF Aβ40 increased in the PIB+ population [56]. These discrepancies might be linked to differences in cohort composition, since the Aβ40 levels may be affected by various pathological conditions [29–31]. The stage of AD, corresponding to various levels of cerebral atrophy probably reducing amyloid production [57], may also account for differences between studies. This is coherent with a recent study confirming the increase of Aβ40 in prodromal AD [58]. Differences in the precision of the analytical methods used, combined with the size of the cohorts, might also explain why only a small, non-significant difference between AD and NAD patients has been observed in some cases.

The present study on Aβ40, which included the largest number of samples studied so far to our knowledge, gave us a sufficiently statistical power to identify small differences. We therefore confirmed the occurrence of an age- and ApoE-independent increase in CSF Aβ40 in AD in comparison with control cohorts consisting mostly of controls and patients with other neurodegenerative diseases and dementia. This observation is valid in different analytical contexts despite differences in the threshold or range for biomarker measurement. It is worth mentioning that in the blood, where the amyloid peptide 42/40 ratio could well indicate the presence of brain amyloidosis [59, 60], it has been established that high Aβ40 levels are associated with greater mortality rate in the elderly [61]. The fact that the CSF and blood amyloid levels are poorly correlated, however, makes it difficult at this stage to extend the present conclusions to this fluid.

The overlap in the CSF Aβ40 values between the AD and NAD populations is worth noting, and the area under the receiver operating characteristic curve (AUC) for AD diagnosis was under 0.8 in all the cohorts tested (SupFigure2). CSF Aβ40 cannot therefore be used as a diagnostic biomarker but could be taken to be a feature “risk factor” in view of the odds ratio of almost 2 recorded on the population having the highest CSF Aβ40 concentration. The increased Aβ40 might be a consequences of a reduced clearance of amyloid peptides in sporadic cases and/or a higher production-lower degradation. This matches the fact that in autosomal dominant forms of AD linked to APP or presenilin mutations [3, 5] and in Down syndrome [4], an overproduction of amyloid peptides is thought to trigger the AD process, along with all its consequences, including tau protein hyperphosphorylation, in particular.

In this context, baseline Aβ40 concentration could indicate subjects with risk of early AD development. The positive correlation found to exist in the present study between Aβ40 and ptau(181) in AD is an additional argument supporting this pathophysiological model. Tau has many phosphorylated isoforms [62, 63], some of them believed to be more specific for AD than p-tau(181), highlighting the pathophysiological role and therapeutic interest of kinases like PKA, CAMkII or Cdk5. This isoform is however one of the best indicator of AD pathology in the CSF where it begins to increase as two decades before the development of aggregated tau pathology [45]. In this work, we had to rely only on the correlation with p-tau(181) because it is the only isoform with in vitro diagnostic (IVD) certification and has been measured in large clinical cohorts. The fact that this correlation was also present in a control population including a subgroup of well-defined FTD [52] and multiple sclerosis patients [51] raises many questions, however. It is tempting to take this relationship to confirm that Aβ peptides may induce the phosphorylation of tau, as observed both *in vitro* and *in vivo* [64]. Since the present study was based on a cross-sectional design, and without neuropathological confirmation, further studies involving a longitudinal design are now required to confirm the idea that high baseline CSF levels of Aβ peptides may have prejudicial effects, leading to AD. The results obtained here are certainly consistent with the idea that approaches making it possible to reduce amyloid beta production rates and levels in the CSF in particular Aβ40, will create valuable new opportunities for developing new curative and/or preventive interventions in AD.

## Data Availability

The datasets used for the analyses are available from the corresponding author on reasonable request.

## Declarations

### Ethics approval and consent to participate

*All the patients at each clinical centre gave their written informed consent to participating in clinical research on CSF biomarkers, which was approved by the respective Ethics Committees. The committee responsible in Montpellier was the regional Ethics Committee of the Montpellier University Hospital and Montpellier CSF-Neurobank #DC-2008-417 at the certified NFS 96-900 CHU resource center BB-0033-00031, www.biobanques.eu. Authorization to handle personal data was granted by the French Data Protection Authority (CNIL) under the number 1709743 v0*.

### Consent for publication

*Not applicable*.

### Availability of data and material

*The datasets used for the analyses are available from the corresponding author on reasonable request*.

### Authors’ contributions

Lehmann, Dumurgier, Lleo, Gabelle had full access to all the data in the study and take responsibility for their presentation and the accuracy of the analysis.

Concept and design: Dumurgier, Gabelle, Lehmann.

Acquisition of the data (biological and clinical): Ayrignac, Marelli, Alcolea, Fortea, Thouvenot,

Delaby, Hirtz, Vialaret, Ginestet, Bouaziz-Amar, Laplanche, Labaug, Paquet.

Drafting of the manuscript: Dumurgier, Gabelle, Lehmann.

Administrative, technical, and material support: Hirtz, Delaby.

### Abbreviations

AD: Alzheimer’s disease
ADNI: Alzheimer’s Disease Neuroimaging Initiative
APP: amyloid precursor protein
AUC: Area under the curve; Aβ: Amyloid β
CAA: cerebral amyloid angiopathy
CI: Confidence interval
CLIA: chemiluminescence immunoassay
CMRR: Centre Mémoire de Ressources et de Recherche
CSF: Cerebrospinal fluid
FTD: Frontotemporal degeneration
FTLD: Frontotemporal lobar degeneration
LBD: Lewy Body dementia
MCI: Mild cognitive impairment
MMSE: Mini Mental State Examination
MRI: magnetic-resonance brain imaging
NAD: Non Alzheimer disease
NFT: neurofibrillary tangles
PET: Positron-Emission Tomography; PiB: Pittsburgh compound B
PLM: Paris-Lille-Montpellier
PSP: Progressive Supra-nuclear Palsy
QC: Quality control
ROC: Receiver operating characteristic
SD: Standard deviation.

